# SHaploseek: A sequencing-only high-resolution implementation of comprehensive preimplantation genetic testing

**DOI:** 10.1101/2023.06.06.23291016

**Authors:** Daniel Backenroth, Gheona Altarescu, Fouad Zahdeh, Tzvia Mann, Omer Murik, Paul Renbaum, Reeval Segel, Sharon Zeligson, Elinor Hakam-Spector, Shai Carmi, David A. Zeevi

**Affiliations:** Braun School of Public Health and Community Medicine, The Hebrew University of Jerusalem, Jerusalem, Israel; PGT Unit, Medical Genetics Institute, Shaare Zedek Medical Center, Jerusalem, Israel; Hebrew University Faculty of Medicine, Jerusalem, Israel; Translational Genomics Lab, Medical Genetics Institute, Shaare Zedek Medical Center, Jerusalem, Israel; Medical Genetics Institute, Shaare Zedek Medical Center, Jerusalem, Israel

## Abstract

**Purpose:** We previously developed Haploseek, a clinically-validated, variant-agnostic comprehensive preimplantation genetic testing (PGT) solution. Haploseek is based on microarray genotyping of the embryo’s parents and relatives, combined with low-pass sequencing of the embryos. Here, to increase throughput and versatility, we aimed to develop a sequencing-only implementation of Haploseek.

**Methods:** We developed SHaploseek, a universal PGT method to determine genome-wide haplotypes of each embryo based on low-pass (≤5x) sequencing of the parents and relative(s) along with ultra-low pass (0.2-0.4x) sequencing of the embryos. We used SHaploseek to analyze five single lymphoblast cells and 31 embryos from 14 families. We validated the genome-wide haplotype predictions against either bulk DNA, Haploseek, or, at focal genomic sites, PCR-based PGT results.

**Results:** SHaploseek achieved >99% concordance with bulk DNA in two families from which single cells were derived from grown-up children. In embryos from 12 PGT families, all of SHaploseek’s focal site haplotype predictions were concordant with clinical PCR-based PGT results. Genome-wide, there was >99% median concordance between Haploseek and SHaploseek’s haplotype predictions. Concordance remained high at all assayed sequencing depths ≥2x, as well as with only 1ng of parental DNA input. In subtelomeric regions, significantly more haplotype predictions were high-confidence in SHaploseek compared to Haploseek.

**Conclusion:** As a single-platform comprehensive PGT solution with higher sensitivity in subtelomeric regions, SHaploseek constitutes a significantly improved, accurate, and cost-effective re-embodiment of Haploseek.

## Introduction

In preimplantation genetic testing (PGT), DNA is extracted from biopsies obtained from *in-vitro* fertilized (IVF) embryos and tested for various molecular variants and chromosomal aberrations. Traditional PGT methods have focused on familial single variants for monogenic (Mendelian) diseases and required family-specific assay preparation. Over the past decade, advances in genome-wide technologies for genotyping and sequencing led to the development of several methods for comprehensive PGT, providing all-in-one solutions for the testing of monogenic disorders, large structural variations, and aneuploidy (PGT-M, PGT-SR, and PGT-A, respectively). Implementations include Karyomapping^1^, OnePGT/haplarithmisis^2–4^, HaploPGT^5^, MARSALA^6^, GENType^7^, FHLA^8^, and others^9–13^. These methods are based on microarray genotyping, genome-wide genotyping by sequencing, whole-genome sequencing, or their combinations. For PGT-M/SR, the data for each embryo must be accompanied by sequencing/genotyping of the embryo’s parents (and, usually, at least one other relative) to determine whether the embryo has inherited the haplotype carrying the pathogenic variant. For a recent review, see reference ^14^.

We previously developed Haploseek^15, 16^, an accurate, comprehensive PGT method designed to be highly affordable. In Haploseek, for PGT-M/SR of inherited variants, the parents and another first degree relative (an already born child, a sample from chorionic villus sampling/amniocentesis, or a grandparent) are genotyped on a microarray. In parallel, embryo biopsies undergo ultra-low-pass sequencing (∼0.4x depth). The genotyping and sequencing data enter a hidden Markov model (HMM), which reconstructs the genome-wide haplotypes of each embryo. The observed sequencing depth at each embryo is used to detect copy number variants. Haploseek was validated as a cost-effective solution for PGT-A, PGT-SR, and PGT-M, including in families of various ethnicities or with consanguinity^15^. Nonetheless, Haploseek uses two different clinical-grade platforms – microarray and next generation sequencing – which is labor intensive and prone to procedural errors. Further, the genome sequencing of the embryos generates data on SNVs not covered by the array, leading to information loss.

Here, we describe the development and clinical validation of SHaploseek, a comprehensive PGT solution based on whole-genome sequencing as the only molecular platform. We validated the accuracy of SHaploseek’s haplotype predictions on a genome-wide scale. We also validated a new low input sequencing protocol for pre-case work-up when DNA is scarce. In subtelomeric genomic regions, SHaploseek has greater diagnostic yield compared to Haploseek. With these improvements, our SHaploseek workflow becomes a reliable universal solution for PGT of most disease-causing variants in the human genome.

## Materials and Methods

### An overview of SHaploseek

SHaploseek is based on low-pass genome sequencing. In addition to the parents, at least one other (first degree) relative must be recruited in order to provide information on the parental haplotypes. The DNA of parents and relatives are sequenced to depth around 1-4x. DNA from the embryo biopsies is amplified and sequenced to very low depth (0.2-0.4x). A hidden Markov model (HMM) uses information from all individuals to infer the parental haplotypes transmitted to each embryo.

### Evaluation of SHaploseek based on cell culture experiments

For our first evaluation of SHaploseek, we used DNA extracted from parents, grown-up children, and cell culture isolates from the same children. We used Families 1 and 2 from our previous work^16^. In each family, we previously performed microarray genotyping of the parents and three or four children. We then designated one child in each family as a phasing reference (referred to as “Child1”) and performed ultra-low-pass (0.2-0.4x) sequencing on whole-genome amplified DNA from single lymphoblast cells derived from each of the other siblings. We combined the array data for the parents and Child1 with the sequencing data for the single cells to reconstruct genome-wide haplotypes for each sibling. Here, for SHaploseek, we replaced the microarray genotyping of the parents and Child1 with low-pass sequencing. We sequenced each of them at just under 9x sequencing depth (Table 1) and then randomly down-sampled each genome to 4x, 2x, or 1x depth for downstream analysis. We inferred the genome-wide haplotypes of each sibling and evaluated their accuracy by comparing them with haplotypes derived from microarray genotypes of bulk DNA from the grown-up children.

**Table 1.** The families who participated in this study and the unsampled sequencing depth of each family member. Parentheses indicate the sequencing depth for individuals who were sequenced a second time based on 1ng of input DNA. N/A, not applicable

| Family | Number of embryos | PGT-M indication | gene | DNA source | Mother depth | Father depth | Child depth | Maternal grandmother depth | Maternal grandfather depth | Paternal grandmother depth | Paternal grandfather depth | Resequencing performed |
| --- | --- | --- | --- | --- | --- | --- | --- | --- | --- | --- | --- | --- |
| 1 | 3 | N/A | N/A | tissue culture | 8.9 | 8.8 | 8.9 | N/A | N/A | N/A | N/A |  |
| 2 | 2 | N/A | N/A | tissue culture | 8.7 | 8.9 | 8.8 | N/A | N/A | N/A | N/A |  |
| 9 | 2 | nonsyndromic hearing loss | GJB2 | whole blood | 13.2 | 13.4 | 8.5 | N/A | N/A | N/A | N/A |  |
| 25 | 3 | HNPCC | MLH1 | whole blood | 22.2 | 13.2 | N/A | 4.9 | 4.8 | N/A | N/A |  |
| 26 | 3 | RCAD | HNF1B | whole blood | 4.9 | 4.9 | N/A | 4.9 | 4.9 | N/A | N/A |  |
| 27 | 4 | 22q microduplication | N/A | whole blood | 4.8 (2.8) | 4.8 (3.4) | N/A | 4.9 (3.2) | N/A | N/A | N/A | V |
| 29 | 2 | t(4;9)(p16.3;q34.3) | N/A | whole blood | 4.8 | 4.9 | N/A | 4.8 | 4.8 | N/A | N/A |  |
| 31 | 3 | Gaucher disease | GBA | whole blood | 4.9 (3.8) | 4.9 (3.2) | N/A | 4.9 (3.3) | N/A | 4.9 (2.8) | N/A | V |
| 33 | 3 | Gorlin syndrome | PTCH1 | whole blood | 13.8 | 22.3 | N/A | N/A | N/A | 4.8 | N/A |  |
| 42 | 3 | Neurofibromatosis | NF1 | whole blood | 4.8 (2.3) | 4.8 (2.3) | 4.8 (3.9) | N/A | N/A | N/A | N/A | V |
| 45 | 3 | Aicardi Goutieres syndrome | SAMHD1 | whole blood | 4.9 (2.8) | 4.9 (5.2) | 4.9 (3.0) | N/A | N/A | N/A | N/A | V |
| 47 | 1 | ADPKD | PKD1 | whole blood | 4.9 (3.1) | 4.8 (1.9) | N/A | N/A | N/A | 4.9 (1.5) | N/A | V |
| 48 | 2 | chr1q21.1 duplication | N/A | whole blood | 8.5 | 8.4 | 8.6 | N/A | N/A | N/A | N/A |  |
| 49 | 2 | Aniridia | PAX6 | whole blood | 8.4 | 8.6 | N/A | N/A | 8.6 | N/A | N/A |  |

### Evaluation of SHaploseek on PGT-M cases

For our second evaluation, we used day 5 blastocyst biopsies that already underwent PGT with both Haploseek^15^ and (for most embryos) classical PCR of informative polymorphic markers surrounding the variant of interest. Among the 12 families, four had an already born child as a phasing reference and eight had one or more grandparents (Table 1). With Haploseek, the parents and other relative(s) were genotyped on microarrays. Here, we sequenced these individuals at depth in the range 5-22x (Table 1), as well as down-sampled to 4x, 2x, and 1x. We inferred the genome-wide haplotypes of each embryo and evaluated the accuracy of our predictions by comparing them to those of Haploseek/PCR.

### Sequencing the parents and reference individuals and variant calling

For both cell culture experiments and PGT-M evaluations, we sent 1 microgram of DNA from the parent and reference individuals to BGI (Hong Kong, China) for genome sequencing. The achieved sequencing depths are listed in Table 1. We aligned the sequencing reads to the human reference genome (hg19) using the Burrows-Wheeler aligner (BWA)^17^. We called single-nucleotide variants using bcftools^18^ in (autosomal+chrX) regions delineated by the gnomAD hg19-v0-wgs_evaluation_regions.v1.interval_list.bed file^19^. These variant calls were only used for initial site filtering, as the phasing method is fully probabilistic (see below). We used bam-readcount^20^ to count reference and alternate alleles at each single-nucleotide variant (SNV) position reported by bcftools. For the down-sampling experiments, we used samtools to down-sample the original alignments to a prespecified mean sequencing depth prior to variant calling and allele counting.

### Whole-genome amplification and low-pass embryo genome sequencing

DNA from either cell culture isolates or blastocyst biopsies was whole-genome amplified (WGA) and sequenced at 0.2-0.4x depth as part of our previous studies^15, 16^. We aligned the sequencing reads to hg19 using BWA and counted reads mapping to reference and alternate alleles using bam-readcount.

For Haploseek, we considered SNV positions matching those on the CytoScan^®^ 750K array (Thermo Fisher). For SHaploseek, we considered, independently in each family, SNV positions identified by bcftools where an alternate allele was present in at least one of the parents or reference individuals. All analyses included both the autosomes and the X chromosome.

### Haplotype prediction for Haploseek

Genome-wide haplotypes for the embryos/cell culture isolates were inferred using an HMM as part of our previous work^15, 16^.

### Haplotype prediction for SHaploseek

Our HMM for haplotype prediction is similar to that we have previously developed for Haploseek^16^. However, for SHaploseek, we modeled the likelihood of observing the sequencing reads (given an assumed genotype configuration) not only from the embryos or single cells (as in Haploseek), but also from the parents and other relatives. We provide full details in the Supplementary Materials and Methods.

### Outputs of SHaploseek

SHaploseek generates two types of outputs. The first is a “binary” haplotype prediction, based on the Viterbi path returned by the HMM. A prediction is provided separately for the maternal and paternal chromosomes of the embryo. Consider the case when the reference relative is a previously born child. In this case, for each SNV and each embryo, the prediction is whether the chromosome is (or is not) identical to that of the reference child. For the case when the reference relative is a grandparent, the prediction is whether the chromosome is coming from that grandparent or from the other grandparent (of the same parent). The second output is the “marginal”, or posterior, probability returned by the HMM. At each SNV, this provides the probability, given the entire data, that the embryo has inherited a haplotype identical to that of the reference child or grandparent. A confident haplotype prediction is reflected by a marginal probability close to 0 or 1. We denote sites with marginal probability >0.99 or <0.01 as high-confidence (“pass”).

### Evaluating the accuracy of SHaploseek

For validating SHaploseek’s single cell or embryo biopsy haplotype predictions, we only considered array sites that had high-confidence haplotype calls in all compared methods. We then dichotomized all marginal probabilities by rounding them to 0 or 1, which generated binary haplotype predictions that could be compared across methods. For the single cell data, we compared the haplotypes predicted by SHaploseek to those predicted from the array genotypes of the corresponding grown-up children^16^. These genotypes are based on bulk DNA and therefore can be considered as “ground-truth”. For the day 5 embryo biopsies, we compared the haplotype predictions of SHaploseek to those of Haploseek. At PGT-M loci, we compared the predictions of SHaploseek to PCR informative polymorphic markers surrounding the variant-based PGT clinical test results. None of the PGT-M loci was directly covered by the array.

### SHaploseek resequencing experiments

For five arbitrarily selected families (Table 1), we repeated the SHaploseek analysis using just 1 ng of input DNA for the parents and reference individuals. We converted the genomic DNA into a genome sequencing library using the Nextera XT library prep kit (Illumina). We normalized and pooled the resultant libraries and then converted them into single stranded circular DNA using the MGIEasy Universal Library Conversion Kit (App –A; MGI) according to the manufacturer’s protocol. We converted the ssDNA library with Illumina adapters into a DNA nanoball sequencing library using the High-throughput Sequencing Primer Kit (App-C; MGI) and loaded the library onto an FCS PE150 flow cell for 2×150 paired end sequencing on the DNBSEQ-G400RS (MGI) high throughput sequencer. We finally used the same embryo data and computational pipeline as described above to predict the haplotypes for the corresponding embryos.

### Visualizations and other statistics

We previously developed a user-friendly web browser interface to visualize Haploseek’s outputs^16^. Here, we updated the interface to accommodate the higher density of SNVs in the sequencing data generated by SHaploseek. We summarized and plotted other data with BoxPlotR^21^ and Statistics Kingdom^22^, online platforms for data analysis and visualization.

## Results

### Replacing microarrays with low-pass sequencing accurately resolves haplotypes in single cells from tissue culture samples

For our initial evaluation of SHaploseek, we compared bulk and single cell data from members of two families, as in our previous studies^15, 16^. In these families, bulk DNA was available from parents and three or four grown-up children each. In our previous experiments, we genotyped the parents and a single reference child from each family using microarrays. We then performed ultra-low-pass genome sequencing of single cells extracted from tissue cultures derived from the remaining children and predicted the haplotypes transmitted to those children. Here, for SHaploseek, we replaced microarray genotyping of the trio (parents and reference child) by genome sequencing at depths ≈9x, and, by down-sampling, 4x, 2x, and 1x. We inferred the haplotypes transmitted to each child based on a hidden Markov model (HMM; Materials and Methods).

We quantified the performance of SHaploseek using two metrics. The first is a measure of the ability of SHaploseek to generate confident output, defined as the proportion of microarray SNV sites where SHaploseek reported high-confidence haplotype predictions. The second is a measure of phasing accuracy, defined as the concordance between the haplotype predictions of SHaploseek, at high-confidence sites, and the haplotypes inferred by microarray genotyping of bulk DNA from the corresponding grown-up children (Materials and Methods).

We report the results in Figure 1, separately for each family, and alongside the performance metrics of the original Haploseek. In Family 1, the proportion of sites with high-confidence haplotype calls, out of ≈200,000 array SNVs, was 89%-99% across methods, children, and sequencing depths (Figure 1a). The haplotype phasing accuracy at these sites exceeded 99.8% under all conditions Figure 1b).

**Figure 1.**
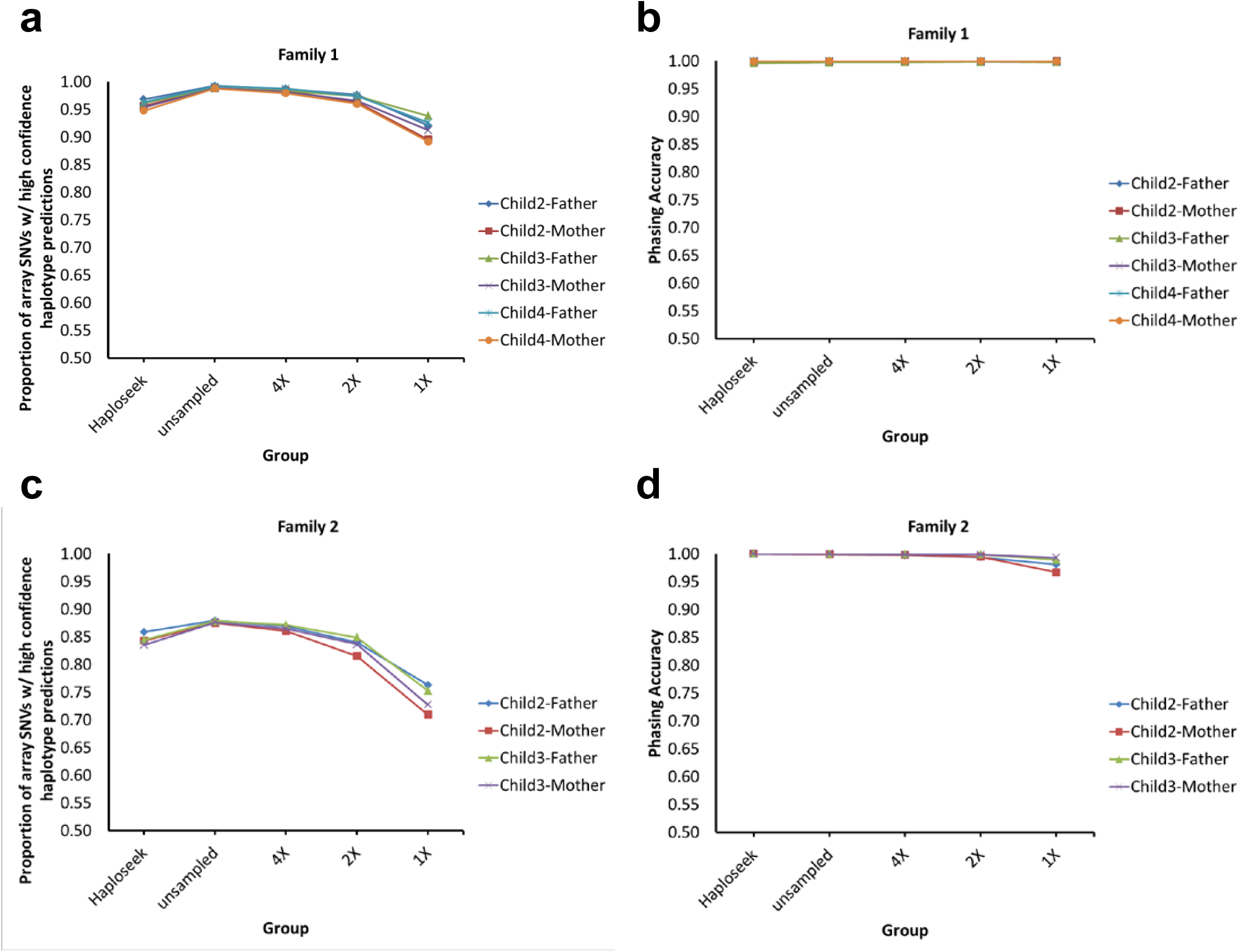
Validation of SHaploseek by sequencing single cells from members of two families. We inferred the haplotypes of grown-up children based on ultra-low-pass genome sequencing of single lymphoblast cells from the children and low-pass sequencing of the parents and a reference child. The “unsampled” sequencing depths of the parent and reference child are given in Table 1. Other sequencing depths were obtained by down-sampling. Haploseek data is also presented for reference. The accuracy of inference of both maternal and paternal transmitted haplotypes is reported for each child (legend). (a) For Family 1, we plot the proportion of the 200,484 genome-wide array SNVs with high-confidence haplotype prediction (marginal probability >0.99 or <0.01) in non-ROC (regions of consanguinity) sites (y-axis). Results are shown for various values of the sequencing depth of the parents and the reference child, as well as for Haploseek (x-axis). (b) The phasing accuracy (based on ‘ground-truth’ haplotypes inferred from bulk DNA) at non-ROC SNVs with high-confidence prediction. (c,d) Same as (a) and (b), for Family 2.

In Family 2, parental consanguinity posed an additional challenge for SHaploseek, as we have previously observed for Haploseek^16^. In what we define as regions of consanguinity (ROCs; regions where the parents share both haplotypes; see ^15^), phasing can be ambiguous, compromising the accuracy of all haplotype phasing methods. Indeed, the presence of multiple ROCs in Family 2 (covering at least 10% of all array SNVs^16^) led to a noticeably lower number of high-confidence sites in that family (71%-88%; Figure 1c). Nonetheless, SHaploseek phasing accuracy in Family 2 non-ROCs exceeded 99.4% at trio sequencing depth of 2x or higher (Figure 1d). At 1x depth, phasing accuracy was compromised, especially in Child 2 (Figure 1c, 1d).

### Clinical validation of SHaploseek with human embryo biopsies from Haploseek PGT cycles

For clinical validation of SHaploseek, we used 12 families in which PGT-M was previously performed with Haploseek (Table 1; Families 9 through 49; Table S1). For each family, we used existing genome-wide sequencing data for the embryo biopsies (one to four embryos per family; depth 0.2-0.4x) and leftover DNA from the embryos’ parents and reference individuals. Four of the families had a previously born child as a phasing reference (ten embryos overall), while the other eight each had one or more grandparents (21 embryos; Table 1; Table S1). We sequenced the parents and reference individuals at depths ≈5-22x, and, by down-sampling, 4x, 2x, and 1x. We evaluated the accuracy of SHaploseek by reporting the proportion of SNVs with high-confidence haplotype prediction in both methods (Figure 2a,c), and, in these SNVs, comparing the genome-wide phasing predictions of SHaploseek to those of Haploseek (Figure 2b,d).

**Figure 2.**
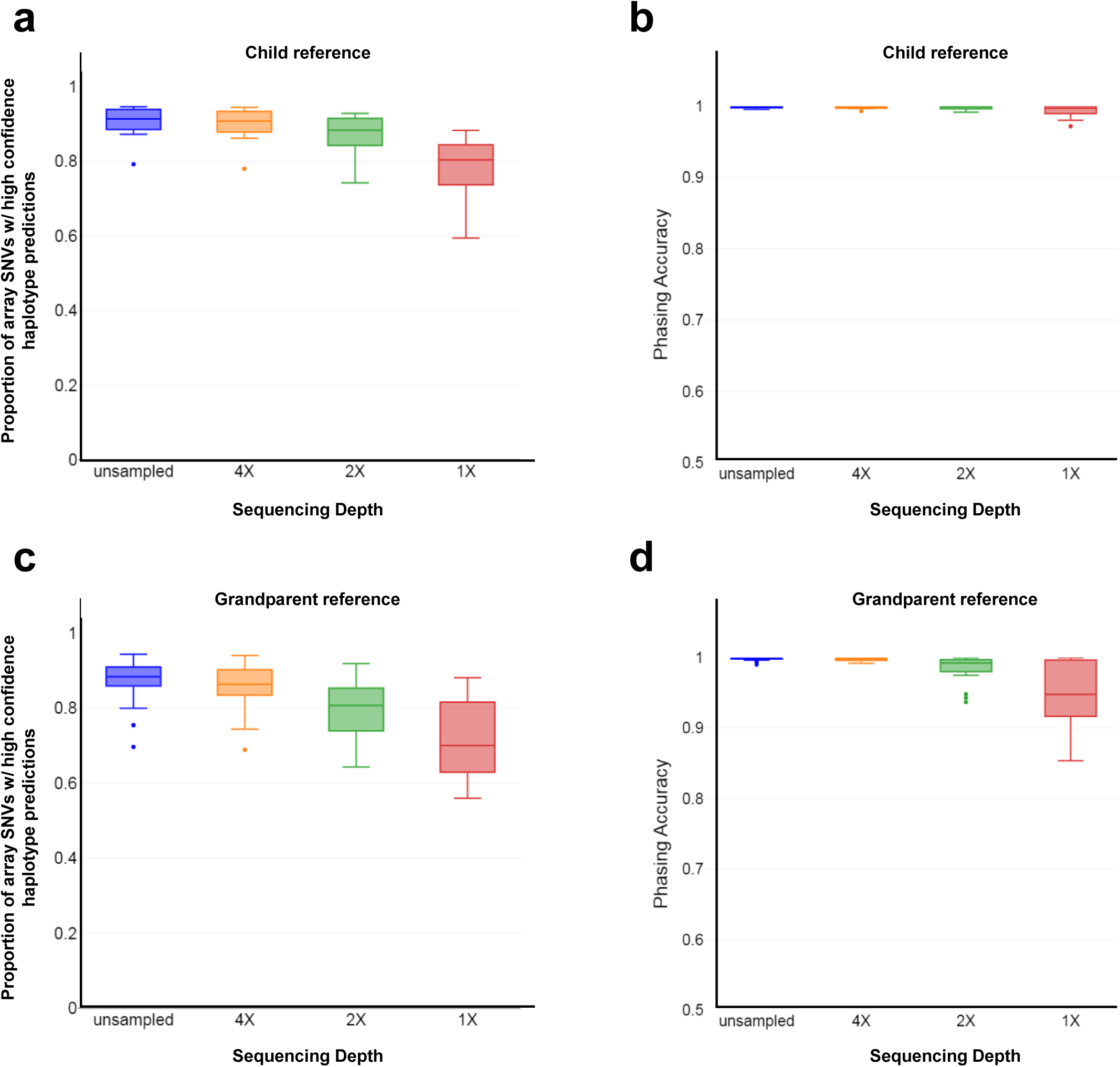
Validation of SHaploseek with clinical PGT embryo biopsies. We evaluated the accuracy of SHaploseek in 31 embryo biopsies from 12 PGT families. (a) In the four families with a child reference individual, we show the proportion of the 200,484 array SNVs with high-confidence haplotype prediction in non-ROC sites in both Haploseek and SHaploseek (box plots). We show results for both the unsampled sequencing data for the parents and reference child (see Table 1 for the sequencing depth of each individual), as well as for lower depths obtained by down-sampling. For each sequencing depth, the box plot represents 20 data points (two for each of the ten embryos), each showing the result of genome-wide prediction of either the maternal or the paternal haplotype of each embryo. (b) Box plots for the haplotype phasing accuracy (measured as the concordance with Haploseek) at SNVs with high-confidence prediction in both Haploseek and SHaploseek, for different sequencing depth categories. (c,d) Same as (a) and (b), respectively, for the eight families with grandparental reference individuals. For each sequencing depth, the box plot represents 24 data points, one for each of the 21 embryos, except the three embryos from Family 31 (Table 1; Table S1) who contributed two data points each, because grandparents from both parents were sequenced.

At high-confidence sites, SHaploseek has very high phasing accuracy (median ≈99%) at all assayed sequencing depths in families with a child reference (Figure 2b), and at sequencing depths ≥2x in grandparent families (Figure 2d). The median proportion of array SNVs with high-confidence haplotype calls was 80%-91% across all sequencing depths in child families (Figure 2a) and 70%-88% in grandparent families (Figure 2c). In grandparent families, the median proportion of high-confidence sites dropped from ≈80% to ≈70% between 2x and 1x sequencing depths (Figure 2c), and was accompanied by a sharp decline in phasing accuracy (Figure 2d).

We next evaluated SHaploseek for the original PGT-M indications, comparing its results against either Haploseek or (for most embryos) also a PCR-based analysis. The evaluation, for various autosomal dominant and recessive disorders, is listed in Table S1. For all sequencing depths, whenever SHaploseek generated a high-confidence haplotype call, it was concordant with the Haploseek/PCR result.

### Clinical validation of SHaploseek after low-pass sequencing of minuscule amounts of parental and reference DNA

Our results suggest that SHaploseek can accurately infer genome-wide haplotypes using low-pass (1-4x depth) parental (and phasing reference) genomes. In the following, we attempted to (i) confirm the accuracy of SHaploseek on low-pass data without the need for down-sampling; and (ii) simulate a clinical case with very low-input, “precious” samples. This can occur, for example, when the reference individual is deceased or when the DNA of a reference child is only available from chorionic villus or amniotic fluid sampling of an aborted fetus. We arbitrarily selected five of the 12 clinical PGT-M families (Table 1) for resequencing at a target depth of 3x based on libraries from just 1ng of input DNA. The DNA samples differed in quality, which led to a wide range of resequencing depths (1.5-5.2x; Table 1). Two of the resequenced families were ‘child’ families (six embryos) and the other three were ‘grandparent’ families (eight embryos). As above, we evaluated the proportion of array SNVs where SHaploseek (and Haploseek) generated high-confidence haplotype predictions, as well as the concordance between the two methods in these sites (Figure 3). The proportion of high-confidence SNVs was 67-87% in the grandparent family embryos and 86%-94% in the child families, and the concordance exceeded 99% in all embryos (99.6% in child families). In PGT-M indication sites, the resequenced SHaploseek predictions were concordant with the Haploseek/PCR-based predictions for all embryos for which SHaploseek generated a high-confidence call (Table S1).

**Figure 3.**
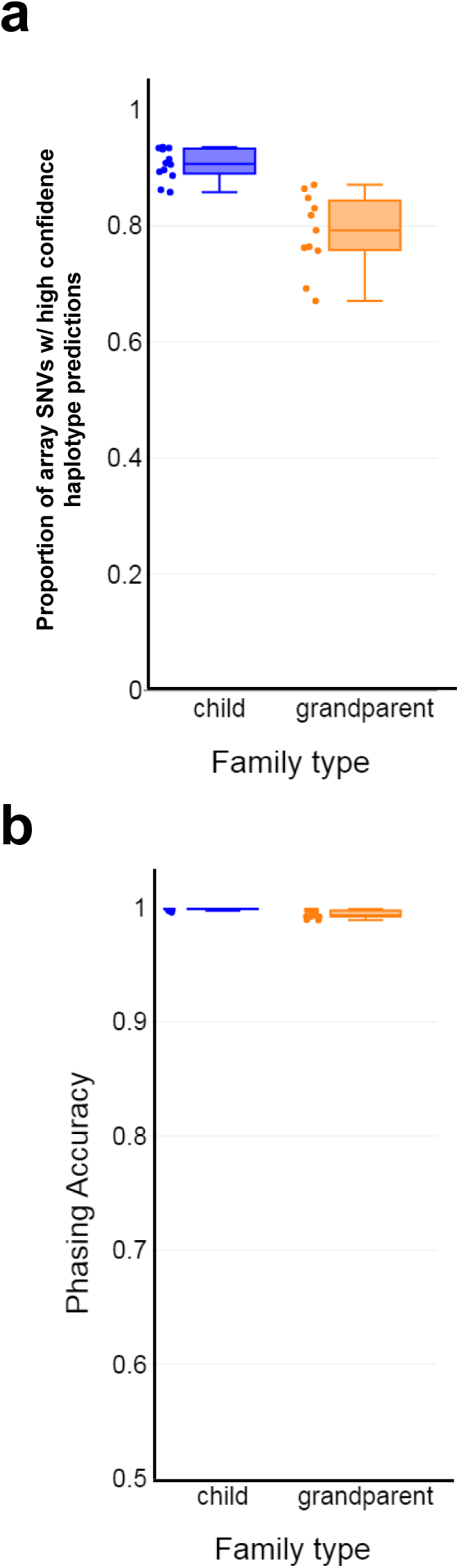
Validation of SHaploseek with clinical PGT embryo biopsies after parental resequencing from low input DNA. We resequenced parent and reference genomes (child or embryo grandparent) at depth 1.5-5.2x from 1ng of input DNA and predicted genome-wide haplotypes with SHaploseek for 14 embryos from five families (Table 1; Table S1). (a) Box plots of the proportion of array SNVs with high-confidence haplotype calls in non-ROC sites in both Haploseek and SHaploseek. The data points include 12 haplotypes for six embryos from ‘child’ families and 11 haplotypes from eight embryos from ‘grandparent’ families. (b) Box plots of the concordance between SHaploseek and Haploseek haplotype calls at high-confidence sites.

### SHaploseek generates higher density and higher quality haplotype predictions than Haploseek in subtelomeric genomic regions

Our experience applying Haploseek to clinical PGT-M cycles was that predictions had low-confidence at SNVs within 5Mb of a telomere or an acrocentric centromere. This is likely due to the small number of flanking SNVs available near the telomeres, which provides insufficient phasing information. The problem is exacerbated in Haploseek, as data is only available for ≈200k SNVs genome-wide. We therefore hypothesized that SHaploseek, which is not limited to a small set of array SNVs, may obtain higher confidence haplotype calls in subtelomeric regions.

To test our hypothesis, we first examined the number of subtelomeric SNVs available for analysis in each method. Indeed, the number of SNVs available for SHaploseek was at least 6-fold higher than Haploseek (Figure 4a). The higher SNV density at subtelomeric regions in SHaploseek translated into significantly higher proportions of high-confidence haplotype calls (Figure 4b). The higher proportions were evident at the unsampled, 4x, and 2x sequencing depths, as well as in the experiments with samples ‘resequenced’ from very low input DNA.

**Figure 4.**
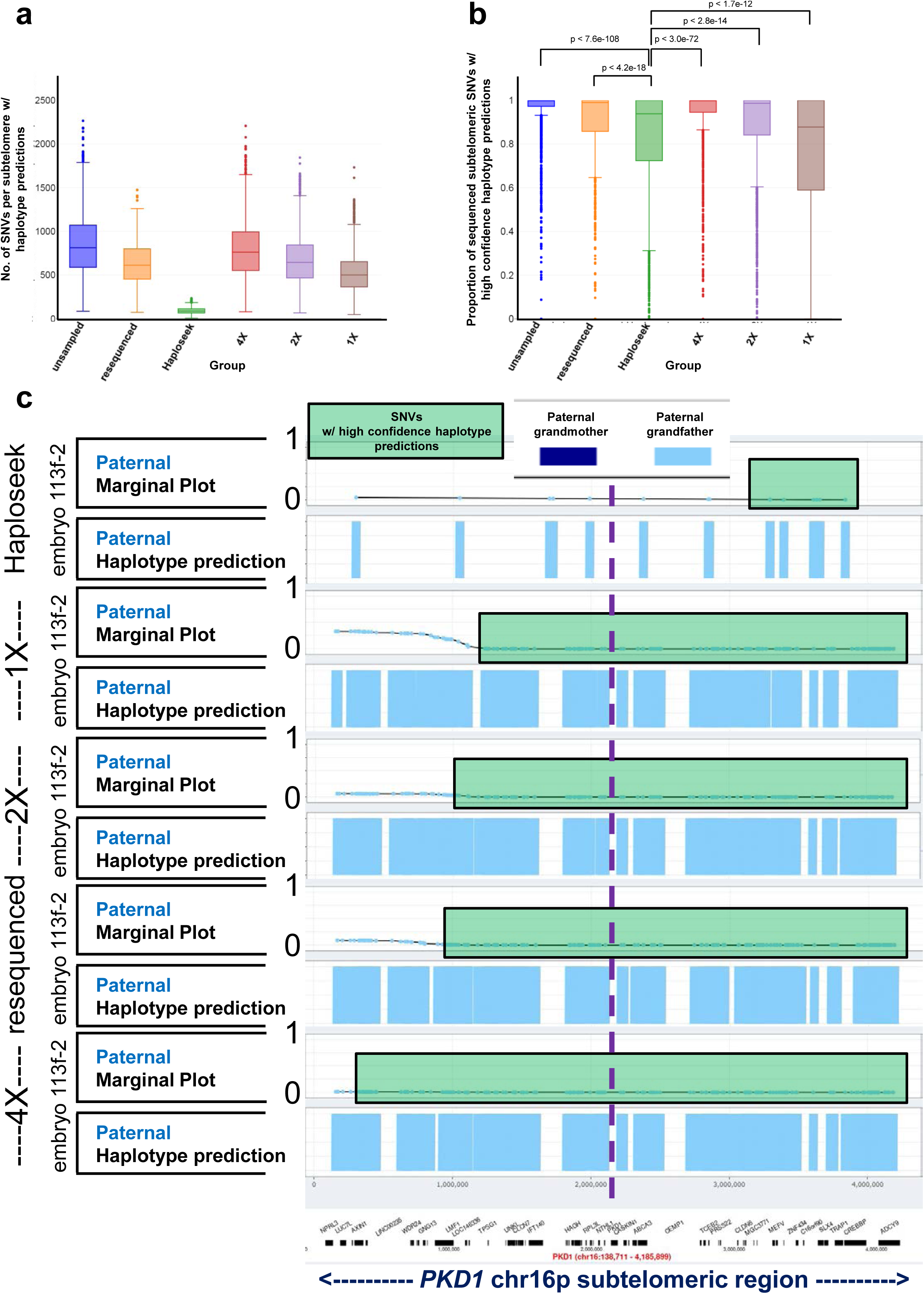
SHaploseek generates higher confidence subtelomeric haplotype predictions than Haploseek. (a) Box plots of the number of subtelomeric SNVs with assigned haplotype predictions. The subtelomeric regions are defined as being within 5Mb distance of an autosomal or chrX telomere or acrocentric centromere. The ‘resequenced’ SHaploseek data set includes 1,038 subtelomeric regions, one on each side of each chromosome, from 23 SHaploseek maternal or paternal haplotypes for the 14 embryos in Figure 3. The remaining categories each include 2,440 subtelomeric regions from 54 Haploseek maternal or paternal haplotypes for the 36 embryos (or single cells) in Figures 1 and 2. (b) Box plots of the proportion of subtelomeric SNVs that were sequenced in the embryo and had high-confidence haplotype prediction in SHaploseek/Haploseek, over the same set of embryos and haplotypes described in (a). We used the Wilcoxon signed-rank test to compare Haploseek with each SHaploseek dataset (p values on top of the plot). (c) Results of Haploseek and SHaploseek paternal haplotype prediction for the *PKD1* gene-flanking subtelomeric 4.2Mb portion of chr16p in embryo 113f-2 of Family 47 (Table S1). For haplotype phasing of the embryo father’s pathogenic variant in *PKD1*, we collected DNA from the paternal grandmother of the embryo (with the same pathogenic *PKD1* variant as the father). In the original Haploseek analysis, we sequenced the 113f-2 embryo biopsy to depth 0.4x and genotyped the parents and the paternal grandmother on arrays. For SHaploseek, we sequenced the parents and grandmother at depths 4.8-4.9x, and down-sampled to 4x, 2x, and 1x. We also resequenced these individuals at depths 1.5-3.1x based on low input DNA (Table 1; see legend on the left of the plot). The paternal haplotypes are depicted in “marginal” and “prediction” plots for each analysis (Materials and Methods). The “marginal”, or posterior, probability indicates the degree of confidence with which the HMM is reporting the haplotype prediction, where a probability of 1 corresponds to certain transmission of the paternal grandmother, and a probability of 0 corresponds to the paternal grandfather. Marginal probabilities <0.01 or >0.99 are considered high-confidence. The marginal probabilities are plotted as light blue dots at SNV sites that were also successfully sequenced in the embryo. SNVs within green shaded rectangles have high-confidence marginal probabilities (here <0.01). The “prediction” plots indicate the HMM binary haplotype predictions (the Viterbi paths), where light blue shaded segments indicate that the embryo haplotype around a given SNV matches that of the paternal grandfather (with the wild type nonpathogenic *PKD1* allele). The approximate location of the *PKD1* gene (chr16:2,138,711-2,185,899; hg19) is marked by a dashed vertical line. Note that the high-confidence region in Haploseek does not encompass the *PKD1* gene and is much smaller than the high-confidence regions of SHaploseek. The wild type (paternal grandfather) *PKD1*-flanking haplotype in embryo 113f-2 was confirmed by PCR-based PGT-M (Table S1).

To illustrate the advantage of SHaploseek in subtelomeric regions over Haploseek, we considered Family 47, with PGT-M indication of a pathogenic variant in the subtelomeric *PKD1* gene on chr16p and a single embryo (113f-2; Figure 4c). In Haploseek, all array SNVs within or flanking *PKD1* had low-confidence calls and thus no result could be reported (Figure 4c). In contrast, both PCR-based PGT-M and SHaploseek (for all considered depths of sequencing) confidently identified a wild type *PKD1*-flanking haplotype (Table S1). The number of subtelomeric SNVs with high-confidence calls in SHaploseek increased with the sequencing depth (Figure 4c).

## Discussion

We described an enhanced, sequencing-only reimplementation of the clinical-grade Haploseek comprehensive PGT method. Our new method, SHaploseek, retains the established user-friendly interface and the universality, affordability, and high accuracy characteristics of Haploseek, along with presenting multiple new advantages. First, it features a single experimental pipeline based on a single platform. Given that sequencing library prep is easily automated, this implies that the hands-on time for family and embryo sequencing is greatly reduced. Second, SHaploseek has higher resolution haplotype predictions, which improves diagnostic sensitivity in difficult subtelomeric regions. Third, the low-pass sequencing in SHaploseek translates into greater sample multiplexing opportunities that would further reduce per-sample costs. Fourth, the waiting time for family haplotype construction can be almost entirely eliminated if the family members are sequenced together with the embryos in the same sequencing run.

In the burgeoning new field of sequencing-based comprehensive PGT solutions, SHaploseek is competitive with recently developed methods. OnePGT^2^ and scGBS^4^, low-pass DNA nanoball sequencing^11^, and GENtype^7^ are all sequencing-based PGT methodologies, like SHaploseek, with the ability to perform PGT-M, PGT-A, and PGT-SR in a single molecular assay. OnePGT, scGBS, and GENtype utilize reduced representation sequencing in order to reduce costs associated with genome-wide haplotype construction^4, 7^. However, the addition of restriction enzyme digestion, size selection, and adapter ligation to the NGS library prep protocol translates into a longer and more laborious (albeit mostly automated) process than what is required for SHaploseek, where DNA fragmentation and adaptor ligation are performed by a transposase in a single 5-minute step. The low pass DNA nanoball sequencing PGT method^11^ is similar to SHaploseek in chemistry, but seems less cost-effective, as it requires sequencing of the parents and embryos at depths 10x and 4x, respectively^11^, compared to ≈3x and ≈0.4x, respectively, with SHaploseek.

Regarding limitations of this study, SHaploseek requires DNA from a family member (beyond the couple) as a phasing reference. While this requirement is common to most comprehensive PGT methods, recent methods such as GENtype and Chen et al.^11^ can work without a phasing reference, provided that the variant of interest can be confidently genotyped in at least one embryo^7, 11^. However, the identification of an embryo bearing the pathogenic variant of interest could require multiple PGT cycles, and may result in misdiagnosis in case of recombination near the variant when only one embryo is available as a phasing reference. Another limitation of the current work is that no follow up was performed on the pregnancy rates and birth outcomes of the embryos assessed.

As mentioned above, one of SHaploseek’s advantages is higher diagnostic yield in subtelomeric regions. In our experience with Haploseek, clinically relevant genes (i.e., *FANCA*, *IKBKG*, *TSC2*, or *PKD1*), submicroscopic deletions, and tandem duplications in these regions were difficult to test, as haplotype calls were often low-confidence, and the risk of embryo misdiagnosis was too high to allow reporting. The increased certainty of SHaploseek in subtelomeric regions is thus a welcome addition to the PGT toolbox. We expect diagnostic yields in these regions to further improve with future versions of SHaploseek, given that repetitive low complexity subtelomeric sequences are now resolved with increasing accuracy by endeavors such as the T2T project^23^.

In our previous work, we predicted that a more convenient, robust, and ubiquitous sequencing-only implementation of Haploseek would not be far away on the horizon^15^. Here we report, in conclusion, that SHaploseek realizes this important goal while retaining the high fidelity and all other positive aspects of the original Haploseek implementation.

## Data availability

Deidentified source data for this study is available upon request.

## Author Information

Conceptualization: D.B., G.A., S.C., D.A.Z.; Data curation: D.A.Z., F.Z., O.M.; Formal Analysis: D.B., G.A., E.H.S., S.Z., D.A.Z.; Funding acquisition: D.A.Z., P.R., S.C., G.A.; Investigation: G.A., D.B., E.H.S, F.Z., O.M. R.S., S.Z., S.C., D.A.Z; Methodology: D.A.Z., D.B, S.C., T.M., F.Z., O.M., S.Z.; Resources: P.R., R.S.,; Software: D.B., F.Z.; Supervision: G.A., E.H.S., P.R., R.S., S.C., D.A.Z.; Validation: D.B., D.A.Z., F.Z., G.A.; Visualization: F.Z.; Writing – original draft: D.A.Z.; Writing – review & editing: G.A., D.B., S.C., D.A.Z.

## Ethics Declaration

Ethical approval for this study was obtained from the Shaare Zedek Medical Center institutional review board. DNA, tissue culture samples, and human embryo biopsies in this study were donated to the Shaare Zedek Medical Genetics Institute for research with informed consent according to Shaare Zedek institutional review board guidelines and as set forth in the Declaration of Helsinki.

## Data Availability

Deidentified source data for this study is available upon request.

